# Transcriptomics of acute DENV-specific CD8+ T cells does not support qualitative differences as drivers of disease severity

**DOI:** 10.1101/2021.09.01.21262833

**Authors:** Alba Grifoni, Hannah Voic, Jose Mateus, Kai Mei Yan Fung, Alice Wang, Grégory Seumois, Aruna D. De Silva, Rashika Tennekon, Sunil Premawansa, Gayani Premawansa, Rashmi Tippalgama, Ananda Wijewickrama, Ashu Chawla, Jason Greenbaum, Bjoern Peters, Vijayanand Pandagrundan, Daniela Weiskopf, Alessandro Sette

**Author notes:** Address correspondence to Alessandro Sette and Alba Grifoni.

## Abstract

While several lines of evidence suggest a protective role of T cells against disease associated with Dengue virus (DENV) infection, their potential contribution to immunopathology in the acute phase of DENV infection remains controversial, and it has been hypothesized that the more severe form of the disease (dengue hemorrhagic fever, DHF) is associated with altered T cell responses. To address this question, we determined the transcriptomic profiles of DENV-specific CD8+T cells in a cohort of 40 hospitalized DENV donors with either a milder form of the disease (dengue fever, DF) or a more severe disease form (dengue hemorrhagic fever, DHF). We found multiple transcriptomic signatures, one associated with DENV-specific Interferon-gamma responding cells, and two other gene signatures, one specifically associated with the acute phase, and the other with the early convalescent phase. Additionally, we found no differences in quantity and quality of DENV-specific CD8+T cells based on disease severity. Taken together with previous findings that did not detect altered DENV-specific CD4 T cell responses, the current analysis argues against alteration in DENV-specific T cell responses as being a correlate of immunopathology.

## Introduction

Dengue disease cases have increased more than 8-fold over the last two decades, reaching 5.2 million infections and 4032 deaths (reported on May-19-2021; www.WHO.org). Dengue disease is caused by infection with the Dengue virus (DENV), a mosquito-borne flavivirus. DENV is endemic in more than 100 countries mainly localized in the tropical and sub-tropical geographical areas and is associated with the potential for even greater spread as a consequence of climate change and global warming (1, 2).

Upon infection, the average incubation period is 4-7 days and followed by a spectrum of disease manifestations ranging from mild febrile symptoms (Dengue Fever, DF) to the more severe clinical symptomatology, characterized by hemorrhagic fever (DHF) and severe plasma leakage, and in the most severe cases hypovolemic shock followed by multi-organ failure (Dengue Shock Syndrome, DSS)(3).

A single Dengue vaccine, CYD-TDV, is currently licensed and indicated solely for subjects previously exposed to one or multiple DENV serotypes (3, 4), thus underlining a need for further improvement of Dengue vaccines. CYD-TDV is based on delivery of the envelope (Env) protein derived from DENV1-4 serotypes, by a Yellow Fever (YF) backbone as a vector (5). Since Env is a strong target for antibody responses, but a weak target for cellular immunity (6-8), vaccine performance might be enhanced by inclusion of other DENV antigens that are dominant target of T cell responses, for example by the use of DENV attenuated viruses as vaccines (5, 9, 10).

In this context, a detailed understanding of the relative role of humoral and cellular immunity in disease protection and immunopathology is key for the design of improved vaccine concepts. Several studies have shown that antibody response can exacerbate disease severity through the phenomenon of antibody-dependent enhancement (ADE), where antibodies generated after primary infection can facilitate viral entry and replication in target cells and trigger a cytokine storm potentially responsible for the leakage observed in severe dengue cases (11-14).

In the case of T cells, it was hypothesized that suboptimal cross-reactive T cells may be unable to control heterotypic infection and rather be associated with an altered phenotype and severe disease (15). This original hypothesis was not confirmed by subsequent studies (16-18). In contrast, several independent studies in the past years have consistently shown a potential contribution of T cells in controlling DENV infection in mice and humans (17, 19-22).

In humans, we and others have shown that in acute infection DENV-specific CD4+T cells are not characterized by changes in phenotype or functional attributes arguing against the notion that altered CD4+ T-cell phenotype or function may be a determinant of severe dengue disease (16, 17, 23). However, the role of CD8+T cells in acute infection and modulation of disease severity remains controversial. To address this point, we compared the transcriptomic profiles of DENV-specific CD8+T cells in a cohort of dengue hospitalized donors with either moderate (DF) or severe (DHF) forms of dengue disease and compared the quantity as well as the quality of CD8+T cell responses.

## Results

### Selection of a dengue hospitalized cohort to investigate the role of CD8+T cells

A total of 40 DENV hospitalized patients were recruited to the study, for the purpose of analyzing the transcriptomic profiles associated of DENV-specific CD8+T cells as a function of DENV disease severity (Table 1). Of those, twenty subjects were associated with hemorrhagic DENV fever (DHF) and twenty were associated with severe DENV fever (DF) with no hemorrhagic manifestations, according to the international WHO criteria prior 2009 (24) and described in more details in the methods.

**Table 1.** General characteristics of hospitalized cohort with Dengue disease.

|  | **Dengue Hemorrhagic Fever**<br>**(DHF; n = 20)** | **Dengue Fever**<br>**(DF; n = 20)** |
| --- | --- | --- |
| **Age (years)** | 14-56 [Median = 33, IQR = 19] | 17-68 [Median = 30, IQR = 18.5] |
| **Gender** |  |  |
| **Male (%)** | 75% (15/20) | 75% (15/20) |
| **Female (%)** | 25% (5/20) | 25% (5/20) |
| **Sample Collection (years)** | 2014-2016 | 2014-2016 |
| **DENV PCR positivity in acute phase** | 25% (5/20) | 20% (4/20) |
| **Antibody test positivity in acute phase** |  |  |
| **IgM (%)** | 80% (16/20) | 75% (15/20) |
| **IgG (%)** | 70% (14/20) | 70% (14/20) |
| **Days Post Fever Onset (PFO)**<br>**at Collection** |  |  |
| **Acute phase** | 1-8 [Median= 5, IQR= 1] | 1-8 [Median= 4, IQR=17 ] |
| **Convalescent phase** | 18-66 [Median= 29 , IQR=1 ] | 18-93 [Median= 30, IQR=12 ] |

The two DHF and DF cohorts were recruited in the 2014-2016 time period and were generally matched in terms of general demographic features (Table 1). The median age of the DF cohort was 33, versus 30 of the DHF cohort, and in both cohorts the female

/male ratio was 25%/75%. At the time of hospitalization 70% of the subjects were DENV IgG seropositive suggesting that the cohorts were predominantly associated with secondary or multiple DENV exposures as previously reported (25). From each subject, a sample was obtained in the acute phase (defined as 1-8 days post fever onset (PFO) and a convalescent phase blood donation, after discharge from the hospital (in the 18 to 93 days range, with median of 29 and 30 days for the DF and DHF cohort, respectively.

### Kinetics of IFN-γ response and memory phenotypes associated with DENV–specific CD8+ T cells

We previously described a DENV CD8 epitope “megapool” (MP) composed of 268 different CD8 T cell epitopes derived from DENV1-4 (6, 26), that allows broad coverage of DENV-specific CD8 responses. Here, PBMCs isolated from blood samples of the DF or DHF cohorts were stimulated *ex vivo* with the DENV CD8 MP at 1 µg/mL. After 3 hours, a cytokine capture assay and flow cytometry analysis were used to isolate IFN-γ+ and IFN-γ-CD8 T cells, as shown in Figure 1A for a representative example.

**Figure 1.**
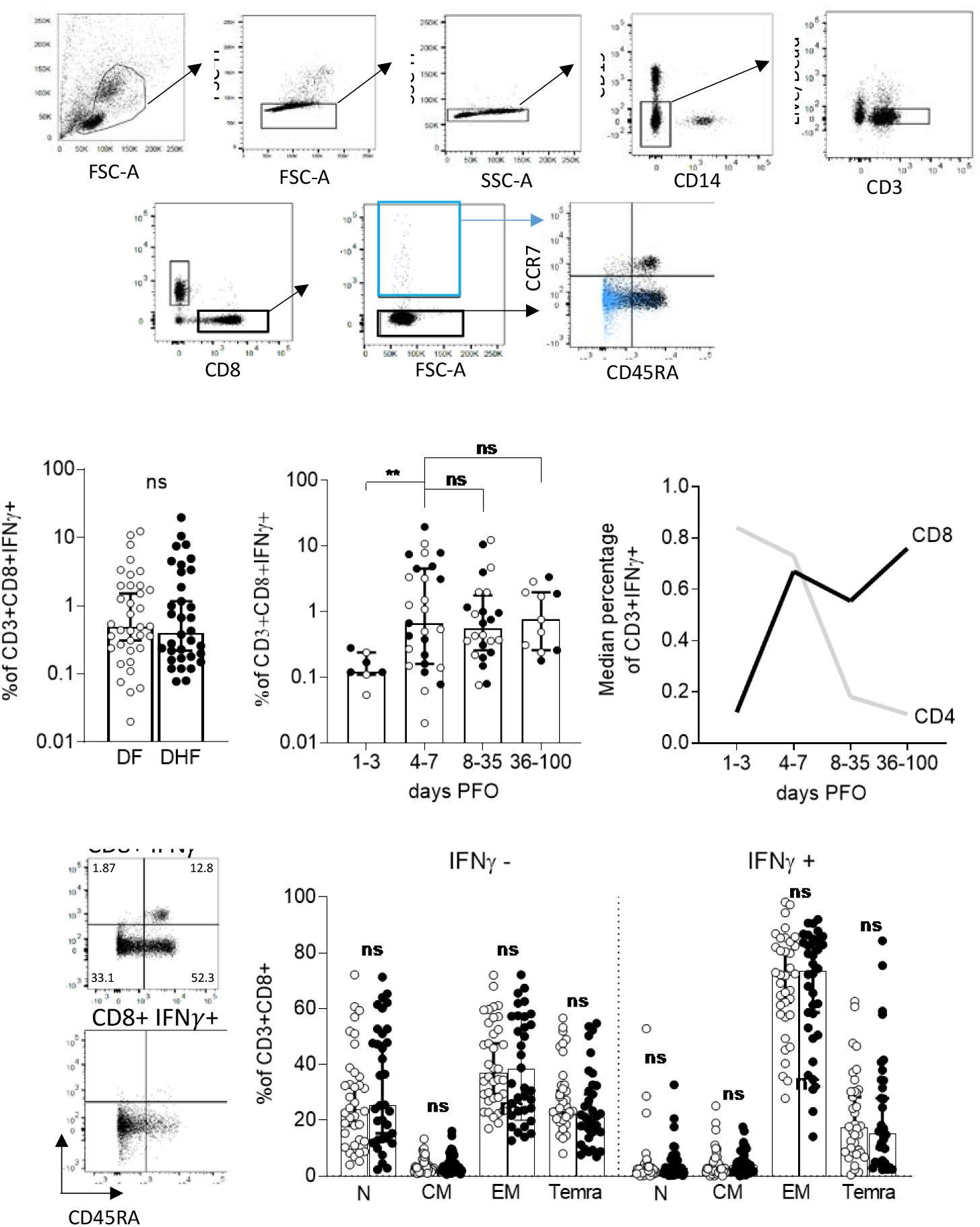
Flow cytometry analysis of DENV-specific CD8+T cells in DENV hospitalized donors. (**A**) Gating strategy. (**B**) Magnitude of CD3+CD8+IFNγ+ T cells responses as a function of disease severity. (**C**) Magnitude of CD3+CD8+IFNγ+ T cells responses as a function of days post fever onset (PFO) (**D**) Median percentage of CD3+CD8+IFNγ+ T cells compared to previous data generated on CD3+CD4+IFNγ+ T cells (3). (**E**) Memory phenotype of CD3+CD8+ T cells in the IFNγ- and IFNγ+ compartments as a function of disease severity. In white, DF donors, in black DHF. ** p<0.01; ns= not significant. Statistical comparisons are performed using Mann-Whitney test. N=naїve; CM=Central Memory; EM=Effector Memory; Temra=Terminal differentiated effector memory cells.

No significant difference was noted in terms of % of IFN-γ producing CD8+ T-cells as a function of DF vs DHF disease severity (Figure 1B; Median±IQR: DF= 0.495±1.978; DHF=0.405±3.2, p= 0.8404 by Mann-Whitney test). Consistent with other reports (27), the CD8+T cell response was low or undetectable in early stages (days 1-3 PFO; Median±IQR=0.12± 0.24), and remained fairly stable thereafter, up to 100 days after viral infection (Median±IQR: 4-7 days PFO=0.67±4.53; 8-35 days PFO= 0.555±1.778; 36-100 days PFO= 0.76±1.97) (Figure 1C).

These kinetics of IFN-γ responses in CD8+T cells are remarkably distinct from those previously reported for CD4+ T cells (17), which peaked during the first week PFO, and decreased thereafter. Our previously published data to CD4+T cell IFN-γ production (17) are replotted here for reference purposes in Figure 1D.

We next determined the memory phenotypes of CD8+ T cells, defined by the expression of the CD45RA and CCR7 cell surface markers. No difference in the memory phenotypes of the DF vs DHF cohorts was observed in either the total CD8+ T cell compartment, or the IFN-γ+ producing CD8+ T cells subset (Figure 1E).

In conclusion, CD8+ IFN-γ+ T cells were detectable from 4 days after PFO onwards and remained at steady levels over a 3-4 months PFO period. No differences were observed as a function of disease severity in the magnitude of IFN-γ+ CD8+T cells responses and their memory phenotypes.

### PCA analysis highlights main sources of transcriptional differences in the study cohort

The results above show that DENV-specific CD8+ IFN-γ+ T cells responses in the DF and DHF cohorts have similar magnitude and kinetics. Next, we investigated potential qualitative differences between the transcriptional profiles of the different T cell populations. To this end, total mRNA was extracted and quantified. We performed micro-scaled RNA-seq assays on the CD8+ IFN-γ+ and CD8+ IFN-γ-T cell populations, sorted after DENV CD8 MP stimulation, as described above and in previous studies (28, 29).

The resulting data was subject to PCA analysis (Figure 2). As expected, a clear separation was observed between IFN-γ+ and IFN-γ-samples along the principal component (PC)1, the main source of variance (PC1 variance=51%; Figure 2A). Two distinct cluster of samples emerged on the PC2 based on the days PFO, which mapped to a first cluster encompassing early (4-7 days PFO) time points, well resolved from a second cluster encompassing later time points (8 days PFO or longer) (PC2 variance=8%; Figure 2B). No clear clustering pattern was observed in samples collected 1-3 days PFO. This is consistent with the kinetics data from Figure 1, which showed that in this time period the IFN-γ response is still developing and of low magnitude. Accordingly, timepoints in the 1-3 days range were excluded from subsequent analyses. Finally, no clear separation in the PCA graph was noted between DF and DHF samples (Figure 2C). In conclusion, these data suggest that while both antigen stimulation and time from infection are associated with clear transcriptomic differences, disease severity is not associated with prominently different transcriptomic profiles.

**Figure 2.**
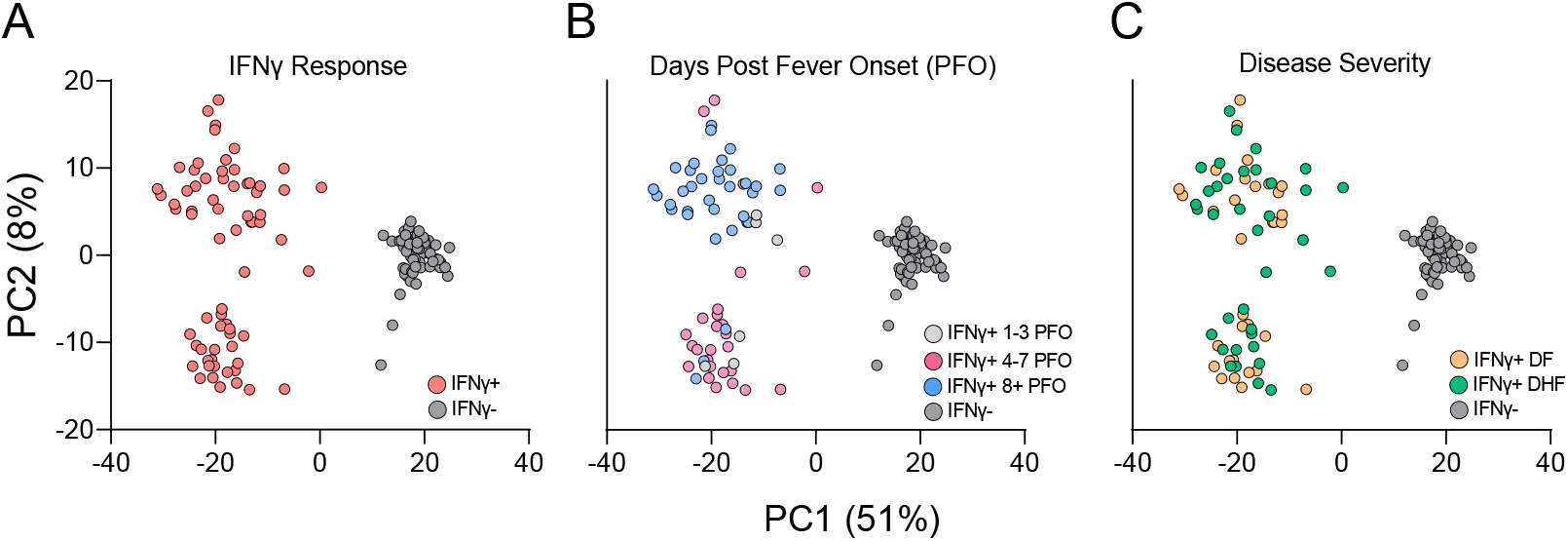
Principal component (PC) analysis of microscaled RNAseq data of DENV-specific IFN-γ+ and IFN-γ-CD8+T cells. PC1 shows separation for IFN-γ- (grey) and IFN-γ+ (red) samples (**A**). PC2 shows separation as function of days post fever onset between 4-7 days (magenta) and 8+ (light blue), no clear separation observed for 1-3 days (light grey) (**B**). No separation is observed as function of disease severity (**C**) DF are shown in orange and DHF in green. Percentage variance explained is shown for PC1 (51%) and PC2 (8%), respectively.

### Characterization of the transcriptional response of DENV-specific CD8+ T cells following cognate antigen stimulation

Based on the PCA analysis shown above, we explored the gene signatures associated with IFN-γ production from DENV-specific CD8+ T cells. When comparing IFN-γ+ CD8+ T cells and IFN-γ-CD8+ T cells, a total of 1806 DE genes were identified, based on Log2 Fold Change (LFC) greater than 1.5 or less than -1.5 and with padj<0.05, and filtering out low-expressed genes using as threshold of exclusion, the median expression level >= 10 tpm across all samples analyzed (IFN-γ-/+) (Figure 3A and Supplemental Table 1).

**Figure 3.**
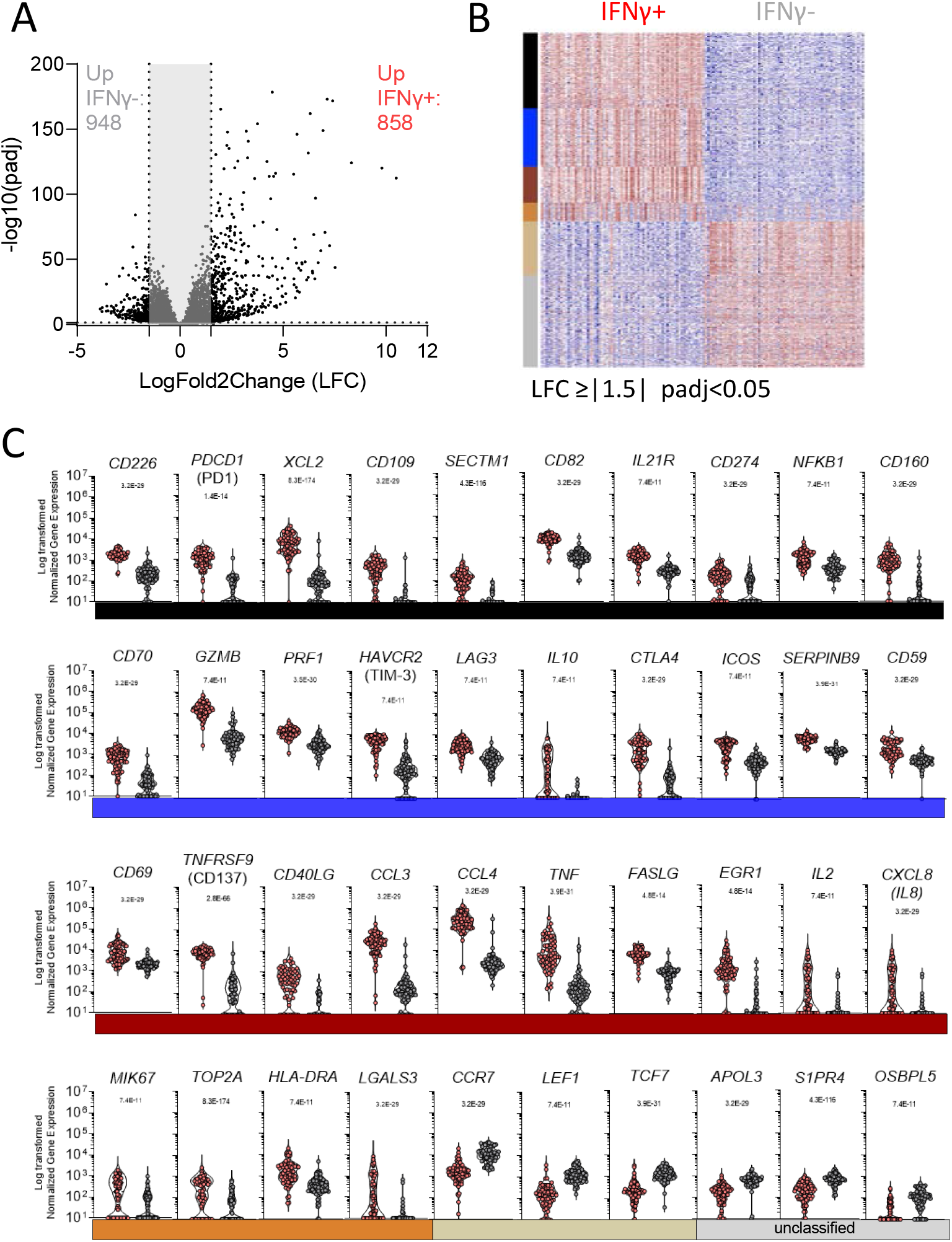
DE genes related to IFNγ signature of DENV-specific CD8+ T cells. (**A**) Volcano plot of differential expressed genes plotted based on their adjusted p values and log2 fold change gene expression. (**B**) Heatmap of the most significantly differentially expressed genes obtained after comparison of IFNγ- and IFNγ+ cells and grouped based on WGCNA analysis. (**C**) Normalized counts of selected genes differentially expressed and related to the WGCNA module/function. All the analyses have been carried considering significant genes with a log2 fold change gene expression less than -1.5 and greater than 1.5 and adjusted p values less than 0.05.

Next, we investigated the function associated with the DE expressed genes by performing a weighted gene co-expression network analysis (WGCNA), to group genes that have similar expression patterns, and identified five main modules (black, blue, orange, tan and coral). Genes that in the network construction did not belong to any specific modules were assigned to a “grey” module. The overall gene expression patterns are shown as a heatmap in Figure 3B ordered by module color (the significant modules and the grouping by color is specified in Supplemental Table 1). The normalized counts observed for the selected genes described above are also visualized in Figure 3C.

The black module included genes related to leukocyte activation and *NFKB*1 signaling, with a specific signature for CTL activation underlined by the joint upregulation of *CD226, CD274* (PD-L1), *CD160* and *PDCD1* (encoding the PD1 protein)(30). The latter was previously associated with DENV-specific CD8+T cells with high cytotoxic potential rather than an exhaustion phenotype (31) (Figure 3C).

The blue module corresponded to additional genes with cytotoxic function, including *CD70, SERPINB9, GZMB* and *PRF1*. The blue module also included genes generally related to regulatory functions, such as *HAVCR2* (encoding TIM-3), *LAG3*, and *CTLA4* (Figure 3C).

The coral module is specifically related to the IFNγ pathway and more in general to cytokine signaling, as shown by the co-expression of CD40LG, *CCL3, TNFA, IL-2* and *CXCL8* (encoding IL-8) (Figure 3C). The orange (peru) module is specifically related to T cell activation in line with the expression of *MKI67 (encoding KI-67)*, generally used as marker of cell activation as well as *HLA-DRA, TOP2A and LGALS3* (encoding gal3) all related to T cell regulation and activation upon stimulation.

The tan module included genes classically related to T cell differentiation, such as *CCR7, LEF1* and *TCF7* all associated with T cell migration and all downregulated in the IFN-γ+ CD8+T cells, which is indicative of a more differentiated phenotype (Figure 3C). Finally, the unclustered genes shown in grey highlight genes related to cell metabolism and particularly to the lipid metabolism such as *APOL3, S1PR4* and *OSBPL5*.

The analyses related to the gene function in each module are summarized in Supplemental Figure 1. In all cases, strong significance was noted for the comparisons, with p values ranging from 7.4E-11 to 8.3E-174.

### Gene signature differences in DENV-specific CD8+IFNγ+ response in early vs late phase samples and as a function of disease severity

In the next series of experiments, we explored the signatures associated with the DENV-specific CD8+IFNγ+ response in early (4-7 days PFO) versus late (8+ days PFO) samples, reflective of acute and convalescent phase samples (Figure 2B). A LFC cutoff greater than 1 and less than -1 was applied in this case, with a more stringent cutoff applied for this comparison to highlight only the most significant genes and the padj <0.05. The data are visualized in a volcano plot (Figure 4A) and in the heat map shown in Figure 4B. A total of 352 genes were differentially expressed, with 167 genes upregulated in the early response and 185 genes upregulated in the late response (Figures 4A and 4B).

**Figure 4.**
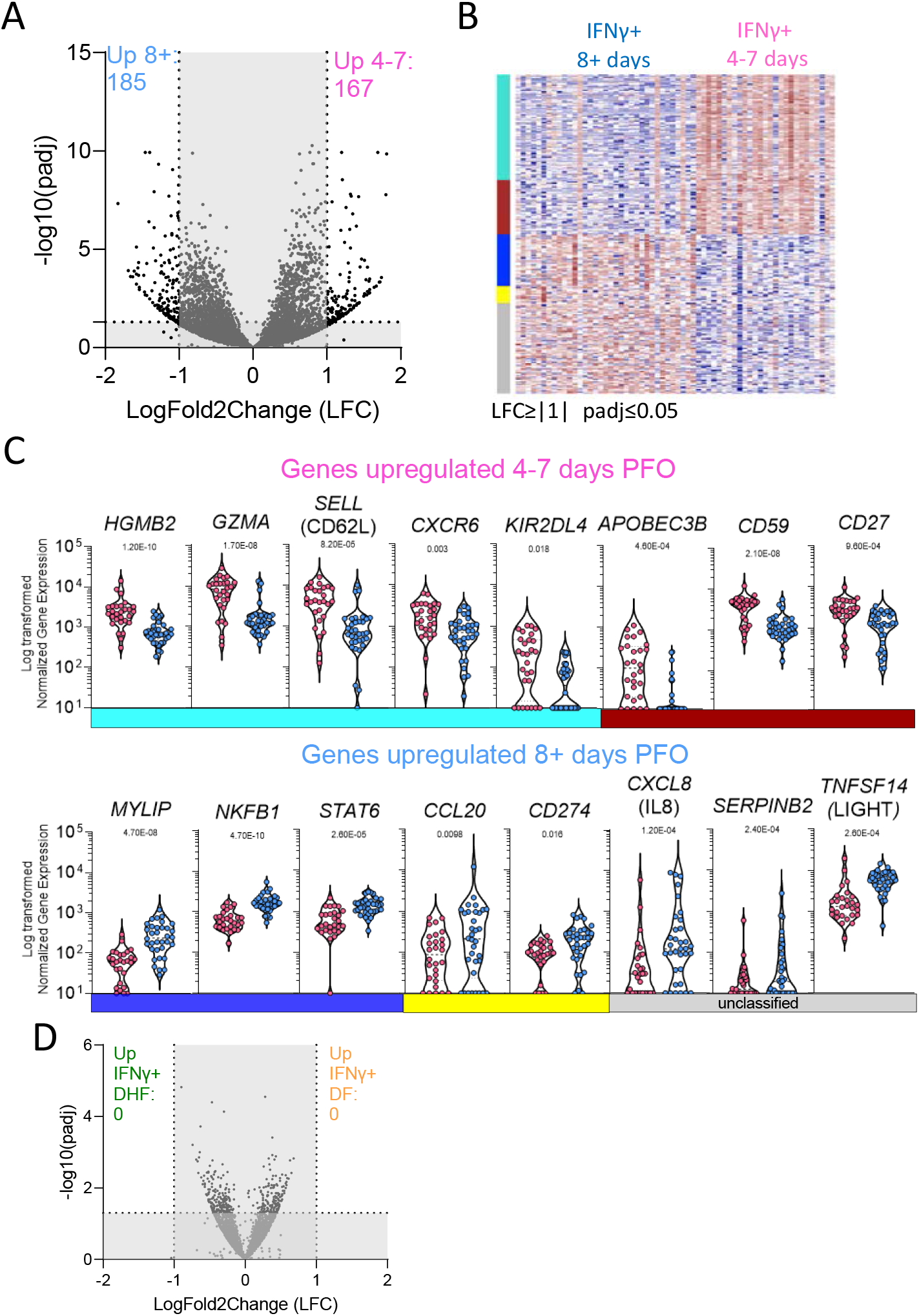
DE genes related to DENV-specific CD8+ IFNγ+ T cells signature. (**A**) Volcano plot of differential expressed genes plotted based on their adjusted p values and log2 fold change gene expression in early and late phase of infection. (**B**) Heatmap of the most Significantly differentially expressed genes in early (4-7 days PFO) and late (8+days PFO) samples. (**C**) Normalized counts of selected genes upregulated in early or late samples. (**D**) Volcano plot of differential expressed genes plotted based on their adjusted p values and log2 fold change gene expression in DF and DHF samples. All the analyses have been carried considering significant genes with a log2 fold change gene expression less than -1 and greater than 1 and adjusted p values less than 0.05.

The WGCNA analysis identified four main modules with group genes that have similar expression patterns (turquoise, coral, blue and yellow) while the genes unclassified were assigned again to a “grey” module. Figure 4B shows the heatmap for this comparison as well as the module color while the significant modules and their function are detailed in Supplemental Table 2 and Supplemental Figure 2. The actual normalized counts observed for those selected genes are also visualized in Figure 4C. Overall p adjusted values ranged from 0.018 to 4.70E-10.

The genes upregulated in the early phase were mostly related to proliferation/cell cycle and HLA class I pathway related such as *HGMB2, CCNB2, KIF15, TOP2A, APOBEC3B* and genes related to the CD8 cytotoxic function such as *GZMA or* cell migration and adhesion such as *SELL* (encoding for CD62L) (Figure 4C). Conversely, genes upregulated in the late phase were not associated with cell proliferation but rather tended to be associated with chemokines pathway such as *CXCL5* and *CXCL8* (encoding for IL8 with a role in angiogenesis) (32) and protein metabolism such as *MYLIP* (encoding for IDOL involved in lipid metabolism) (33) and *SERPINB9* characteristic of a more quiescent cell cycle (34) (Figure 4C).

Finally, we investigated whether any differential gene signature could be discerned comparing the DF and DHF samples. As described above, DE genes were defined by LFC greater than 1.5 and padj <0.05 and the volcano plot is shown in Figure 4D. Consistent with the exploratory PCA analysis shown in Figure 2C, no genes were found to be differentially expressed. In conclusion, these data show that a differential gene expression profile is associated with the early vs late phase of the response, while the transcriptomic profile associated with DENV-specific CD8+ T cells in DF versus DHF samples is remarkably similar.

## Discussion

Herein we reported that specific transcriptomic profiles associated with DENV-specific CD8+ T cells, and in the acute vs recently convalescent phase. We further showed the transcriptomic profiles associated with different disease severities (DF vs DHF) are remarkably similar.

In previous studies, we characterized DENV-specific CD4+ and CD8+T cells responses in the general population of endemic areas (6, 7, 17, 35-37). The results enabled the generation of pools of experimentally defined epitopes with broad HLA coverage spectrum and suited to analyze antigen specific T cell responses in small-volume samples (38). The current results further exemplify how the MP approach is well suited to obtain granular characterization of antigen specific T cell responses.

These results shown are significant in several different respects. First, they define the signature associated with DENV-specific CD8+ T cells following actual activation with cognate antigen. The transcriptomic patterns associated with DENV-specific CD8+T cells revealed three different gene signatures. The first signature, characteristic of the acute disease stage and associated with T cell proliferation and HLA-specific activation; the second signature characteristic of transition to memory phenotype and observed in the post febrile and early convalescent stage; and a third stable signature observed in both acute and convalescent phases, related to sustained cytokines release and T cell activation. Thus, the present study details the patterns of antigen specific CD8+ T cell gene activation associated with acute DENV disease to an unprecedented level of granularity.

Our study defines, as mentioned above, the patterns of gene expression of DENV specific T cells. This is important, to rule out non-antigen specific effects associated with bystander activation. In this context, Chandele and co-authors measured T cell activation in a Thailand cohort and showed an increase of HLA-DR and CD38 markers in the DHF/DSS population in absence of an equivalent increase of IFNγ (39). This result is consistent with a bystander activation effect driven by the pro-inflammatory environment characteristic of the severe dengue disease. The study of Waickman et al., of healthy individuals after TAK-003 vaccination, another dengue vaccine based on delivery of the envelope (Env) protein derived from DENV1-4 serotypes in a DENV2 backbone, indeed showed that HLA-DR and CD38 markers also identified a large amount of TCR clones unrelated to Dengue-specific specificity (40).

Our study is complementary to the recent study of DENV-specific CD8+T cells restricted on three specific HLA molecules (HLA-B*58:01, HLA-A*01:01 and HLA* 24:02) (16) identified by tetramer staining, where the cells were characterized in absence of cognate antigen stimulation. Chng *et al* found two major populations in convalescent T cells characterized by the expression of CD57 and CD127 markers stable up to one year post dengue infection. Here, while we did not observe a cluster of cell populations in our convalescent samples, we did observe other markers consistent with memory differentiation such as downregulation of TCF7, CCR7 and LEF1. Additionally, consistent with Chng *et al*., we detected upregulation of classical activation genes MKI-67, *HLA-DR* and *CD38* genes as well as *CD69, ICOS* and *PDCD1* (encoding PD1). Taken together these results underline how different approaches (tetramer based and DENV-epitope stimulation) generate complementary insights in the process of activation of DENV - specific T cells.

The present analysis is consistent with the activated gene signature we previously reported for DENV-specific CD8+ T cells collected from blood bank healthy DENV-seropositive donors, and previous studies in acute samples by Chandele et al., (37, 39). In particular, our previous study identified genes DE in DENV specific Tem vs Temra CD8+ subsets utilizing PBMC samples from the Colombo (Sri Lanka) blood bank, which are therefore akin to the convalescent samples studied herein (41). Despite the differences in experimental design (the Tian et al., 2019 study was designed to study DE genes in different memory subsets) several genes that were identified in that study were also identified in the current analysis, including *CCL3, FOSL1, TNF, XCL1* and *CD160* as shown in Supplemental Table 1. It is worth noting that in this analyses we also found expression of *TNF*SF*9* (encoding CD137/41BB) and *CD69* markers, highlighting the potentiality to use the combination of these markers to identify DENV-specific CD8+T cells and their functionality, as recently reported in the context of SARS-CoV-2 (42).

Similar observations were made at the level of DENV-specific CD4+ T cells, where the acute phase of infection was associated with a subset of CD4+ T cells that co-produced IL-10 and IFN-γ, but no altered phenotype was associated with DHFs (17). Importantly, the analysis of DENV-specific CD4+T cells in endemic areas of healthy subjects and severe patients also did not shown different or altered phenotypes as correlating with disease severity (17, 43).

Both DENV-specific CD4+ and CD8+ T cells during acute phase of the disease expressed multiple activation and inhibitory genes such as *CTLA4, ICOS, LAG-3, HAVCR2* (encoding TIM-3) and *IL-10* as well as effector molecules *GZMB* (encodes Granzyme B), and *CCL4*, suggesting that cytotoxic activity is exerted by both populations during acute phase of infection (30, 43, 44).

In this study, we observed a difference in the kinetics observed for the DENV-specific T cells, with a decrease in DENV-specific CD4+T cells right after acute phase of infection (4-7 days post fever onset, PFO) and a more stable DENV-specific CD8+ T cells up to 100 days PFO. This difference in kinetics is also shown in the Chng study, highlighting the importance of DENV-specific CD8+T cells in long-term protection (16). Of particular relevance to our understanding of DENV protective immunity and DENV-associated immunopathology, we found no association with either quantitative or qualitive differences of the DENV-specific CD8+ T cell responses with disease severity. Taken together with previous studies (23, 45, 46), it is possible to speculate that the overall DENV-specific T cell responses play an important role in viral clearance (23), while the increased pro-inflammatory environment, bystander activation and dysfunctional innate immune responses maybe associated with immunopathology (47). Indeed, previous studies have shown that dengue-infected monocytes signatures differ as a function of disease severity (48). In conclusion, the lack of qualitative differences of DENV-specific CD8+ and CD4+ T cells as a function of disease severity does not support a pathogenic role of T cells in the severity of dengue infection.

## Methods

### Human Blood Samples

Human blood samples were collected in the North Colombo Teaching Hospital, Ragama in Gampaha District, Sri Lanka and in the National Institute of Infectious Diseases, Gothatuwa, Angoda, Sri Lanka. PBMC were isolated from small blood volumes from anonymous patients at the time of diagnosis/admission and at discharge from the hospital or after recovery from the disease. Patients were diagnosed either as dengue fever (DF), having no sign of bleeding or pleural effusion, or as dengue hemorrhagic fever (DHF). Serology was confirmed by DENV specific IgG ELISAs and flow cytometry-based neutralization assays as previously described (49).

### Peptides and Megapools

To stimulate DENV specific CD8^+^ T cells, 268 DENV-specific CD8 epitopes that account for 90% of the IFNγ response in both Sri Lankan and Nicaraguan cohorts (6, 26, 30) were synthesized (A&A, San Diego, CA), resuspended in DMSO, pooled, lyophilized, and resuspended as previously reported (38) at 1mg/ml.

### IFN-γ capture assay and cell sorting for RNA-Seq

Human PBMC were thawed and rested overnight at 37° C in HR5 culture media consisting of RPMI (catalog#RP-21, Omega Scientific, Inc.), Human serum at 5% (catalog# 100-512, Gemini Bio-Products), 2mM l-alanyl-l-glutamine (GlutaMAX-I, catalog# 35050061, Thermo Fisher Scientific), and 100 U/ml penicillin, andB 100 μg/ml streptomycin (catalog# 400-109, Gemini Bio-Products). After overnight resting cells were stimulated with DENV CD8 megapool (1ug/mL final concentration) for 3 hours at 37°C.. IFNγ producing cells were labelled using IFN-γ Secretion Assay – Detection kit (PE) (catalog#130-054-202, Miltenyi Biotec) according to manufacturer’s instructions. Cells were additionally stained with anti-CD3 (AF700, clone OKT3, catalog#317340 Biolegend), CD4 (APC eFluor 780, clone RPA-T4 catalog# 301042, eBioscience) CD8 (BV650, clone RPA-T8 catalog# 47-0049-42, Biolegend), CD14 (V500, clone M5E2, catalog# 561391, BD Biosciences), CD19 (V500, clone HIB19, catalog# 561121, BD Biosciences), CD45RA (eFluor 450, clone HI100, catalog# 48-0458-42, eBioscience) CCR7 (PerCP Cy5.5, clone G043H7, catalog# 353220, Biolegend), and viability dye (ef506, catalog# 65-0866-18, eBioscience). The CD8+ T cell population was isolated according to the gating strategy detailed in Figure 1A. 200 cells each from CD8+IFN γ+ and CD8+IFNγ-cell populations were sorted using a FACSAria cell sorter (BD Biosciences). Data was analyzed using Flowjo version 10.6 (Treestar Inc.). Statistical analysis and visualization were performed using Prism8.4.3 (GraphPad).

### Microscaled RNA-Sequencing assay

200 to 400 sorted cells were directly collected in 0.2 mL pcr tubes (Axygen) containing 8µL of lysis buffer (0.2% Triton X-100 [catalog#X100-100mL, Sigma-Aldrich], 0.2 mM dNTP [catalog#R0193, ThermoFisher Scientific], 1U/uL recombinant RNAse Inhibitor [catalog#2313A, Takara Bio]). Once cell collected, tubes were vortexed, spun down, and stored at -80° C. Microscaled RNA-Seq was performed according to the Smart-Seq2 protocol as previously reported (28, 50). Briefly, mRNA was captured using poly-dT oligos an directly reverse-transcribed into full-length cDNA using the template-switching oligo. cDNA was amplified by PCR for 19 cycles and purified using the AMPure XP magnetic bead (0.9:1 v/v ratio, Beckman Coulter). 1ng of cDNA from each sample was used to prepare dual-index barcoded standard NextEra XT Library (NextEra XT DNA library prep kit and index kits, Illumina). Both whole transcriptome amplification and sequencing library preparations were performed in a 96-well format to reduce assay-to-assay variability. Quality control steps were included to determine the optimal number of PCR preamplification cycles, and library fragment size. Samples that failed quality controls were eliminated from downstream steps. Libraries that passed quality controls were pooled at equimolar concentration, loaded, and sequenced on the HiSeq 2500 (Illumina). Libraries were sequenced to obtain more than 8 million 50-bp single-end reads (HiSeq Rapid Run Cluster and SBS Kit v2, Illumina) mapping uniquely to mRNA reference, generating a total of about 204.3 million mapped reads (median of about 8.6 million filtered mapped reads per sample) libraries.

### RNA-Seq analysis

The paired-end reads that passed Illumina filters were filtered for reads aligning to tRNA, rRNA, adapter sequences, and spike-in controls. The reads were then aligned to GRCh38 reference genome and Gencode v27 annotations using STAR (v2.6.1)(51). DUST scores were calculated with PRINSEQ lite v0.20.3(52), and low-complexity reads (DUST > 4) were removed from the BAM files. The alignment results were parsed via SAMtools (53) to generate SAM files. Read counts to each genomic feature were obtained with the featureCounts (v 1.6.5)(54) using the default option along with a minimum quality cut off (Phred > 10). After removal of absent features (zero counts in all samples), the raw counts were then imported to DESeq2 v1.28.1 to identify the differentially expressed genes between groups(55). P-values for differential expression are calculated using the Wald test for differences between the base means of two conditions. These P-values are then adjusted for multiple test correction using Benjamini Hochberg algorithm (56). Genes with an adjusted P value less than or equal to 0.05 and an absolute shrunken log2 fold change greater than 1.5 were considered differentially expressed between CD8+IFNγ+ and CD8+IFNγ-groups. Genes with an adjusted P value less than or equal to 0.05 and an absolute shrunken log2 fold change greater than 1 were considered differentially expressed between CD8+IFNγ+ 4-7 days from fever onset and CD8+IFNγ+ 8+ days from fever onset groups. Principal Component Analysis (PCA) was performed using the ‘prcomp’ function in R. The sequences used in this article have been submitted to the Gene Expression Omnibus under accession number GSE174482 (http://www.ncbi.nlm.nih.gov/geo/).

### WGCNA analysis

Weighted gene co-expression network analysis (R package WGCNA v1.69) was performed for genes with median DESeq2 normalized counts greater than 10 in either of the 2 groups IFNγ- or IFNγ+(57, 58), a p adjusted value of <= 0.05 and an absolute foldchange cut off of 1.5 in the DE analysis between IFNγ+ and IFNγ-samples. A total of 5 coexpression modules across 992 genes were identified, the genes that were not assigned to any of these modules were kept in a separate gray module. The genes that were assigned to a module but had a negative module membership value (genes with low intramodular connectivity) were also assigned to the gray module. The gene expression profile of a module was summarized by module eigengene, which is defined as the principal component of the module. T tests were performed to compare module eigengene values between samples from each of the two given groups and all of the modules were found to have significantly different gene expression profiles between the two groups. A row-scaled heatmap of log-transformed normalized counts of these DE genes was created with the ‘heatmap.2’ function in gplots library in R v4.0.3. The modules were used as RowSideColors. The gene function was inferred by using Metascape(59).

### Data Availability

The RNA-Seq data were deposited in the NCBI’s Gene Expression Omnibus (GEO) database under the accession code GSE174482 and ImmPort under the study number SDY888.

### Study approval

The institutional review boards of both the La Jolla Institute for Immunology and the Medical Faculty, University of Colombo (serving as the NIH-approved Institutional Review Board for Genetech and Kotelawala Defence University), approved all protocols (VD-064, VD-085, EC-15-095) described in this study.

## Data Availability

The RNA-Seq data were deposited in the NCBI Gene Expression Omnibus (GEO) database under the accession code GSE174482

## Acknowledgments

Research reported in this publication was supported by the National Institute of Allergy And Infectious Diseases of the National Institutes of Health under Award Number U19 AI118626 to A.S and B.P. In addition, the HiSeq 2500 was acquired through the NIH Shared Instrumentation Grant (SIG) Program (S10): S10OD016262. We would like to thank LJI Flow cytometry Core for their support. The FACSAria II Cell Sorter was acquired through the Shared Instrumentation Grant (SIG) Program (S10) S10 RR027366.

## Authorship contributions

AG, DW, AS designed the experiments and wrote the manuscript. AG, HV and JM performed the cell experiments. Alice W, GS performed the RNA sequencing experiments under VP supervision. HV, KMYF and AC performed bioinformatic analyses under the supervision of BP and JG. ADDS, SP, GP, Rashika T, Rashmi T and Ananda W were involved in sample collection, sample processing and collection of clinical information. AS secured resources.

## Supplemental Figures

**Supplemental Figure 1.**
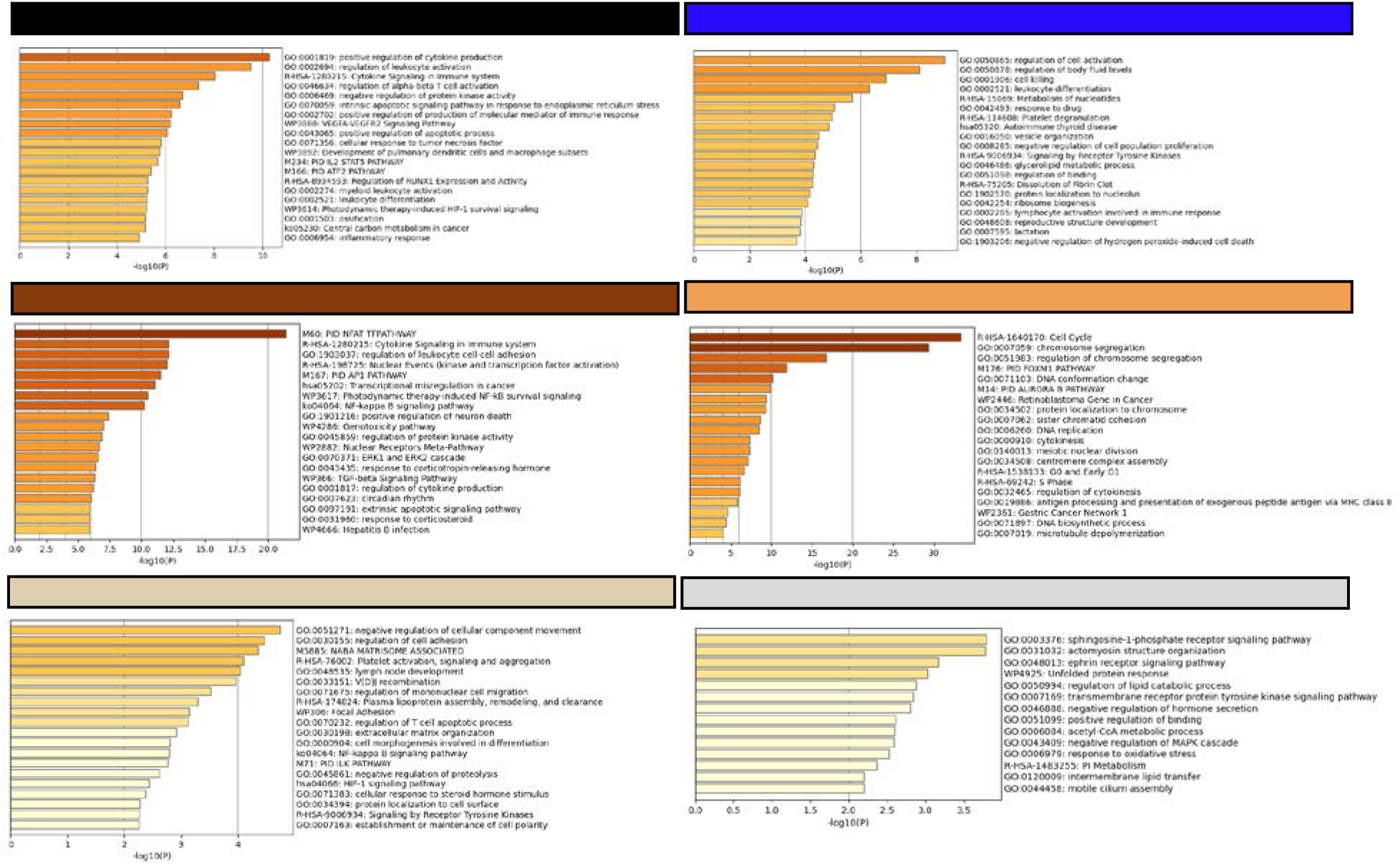
Functional annotation of WGCNA modules based on DENV IFNγ+ versus DENV IFNγ-comparison. The list of DE genes as a function of the modular analysis shown in Figure 3 have been separately investigate to associate a functional annotation per each of the significant modules in analysis. The gene function was inferred using Metascape (http://metascape.org).

**Supplemental Figure 2.**
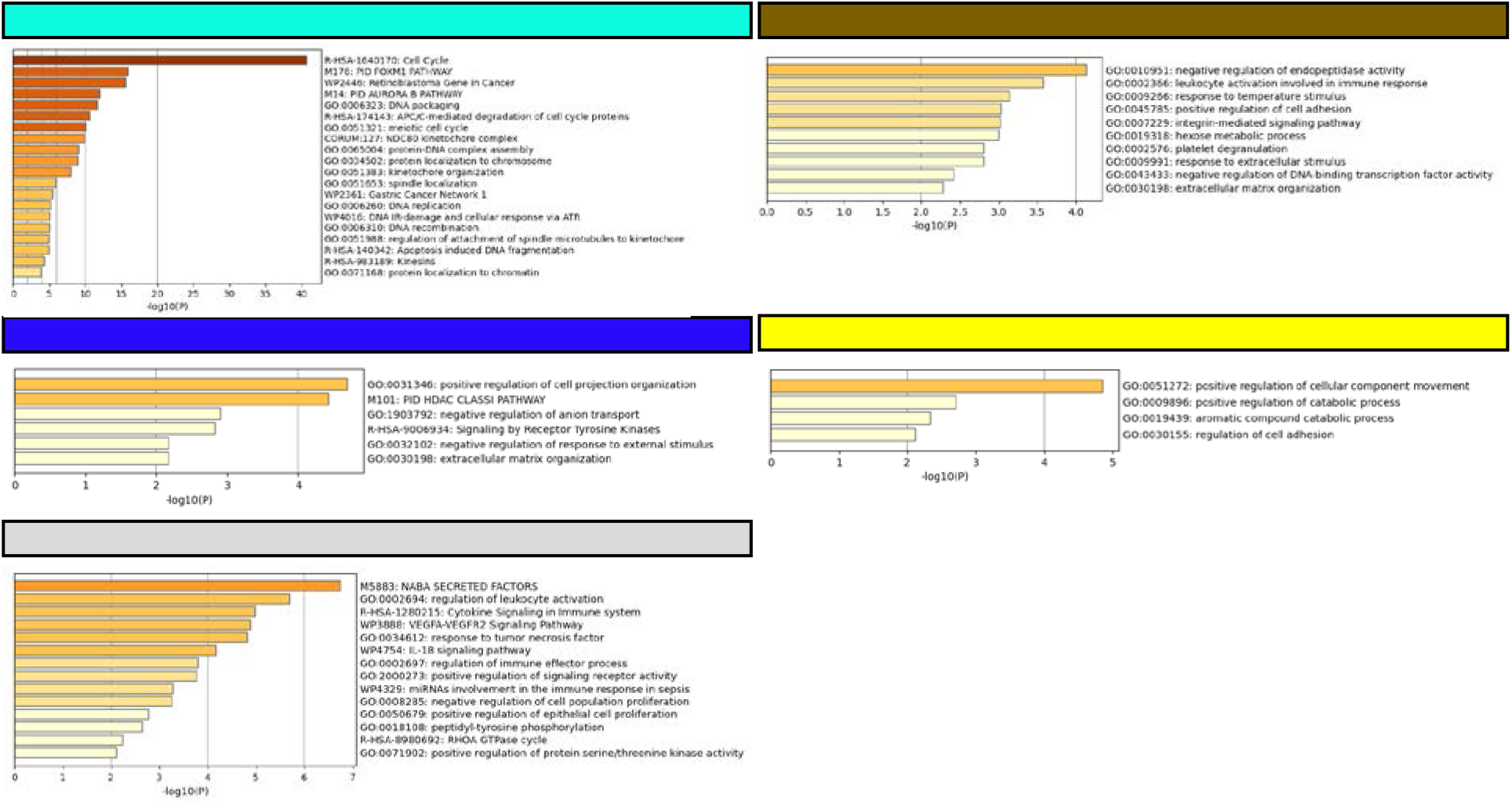
Functional annotation of WGCNA modules based on DENV IFNγ+ samples collected 4-7 days versus 8 or more days post infection. The list of DE genes as a function of the modular analysis shown in Figure 4 have been separately investigate to associate a functional annotation per each of the significant modules in analysis. The gene function was inferred using Metascape (http://metascape.org).

